# COVID-19 and Herpes Simplex Virus Infection: A Cross-Sectional Study

**DOI:** 10.1101/2021.07.09.21260217

**Authors:** Mohammed Shanshal, Hayder Saad Ahmed

**Author notes:** **Corresponding author:** Mohammed Shanshal, MBChB, FABMS – Dermatology, Specialist Dermatologist at Basildon University Hospital, UK, **Email:**. Specialist dermatologist, MBChB, FABMS – Dermatology (Mohammed Shanshal). Specialist dermatologist, MBChB, FIBMS – Dermatology (Hayder Saad Ahmed). **Conflicts of interest:** None. **Funding sources:** None. **Patient privacy and ethical use of images:** The patients in this manuscript have provided written informed consent for the publication of their case details, including the use of images.

## Abstract

**Background:** Despite being variable and poorly characterized, the reported cutaneous manifestations of COVID-19 are of increasing concern.

**Aim of the Study:** This study aimed to assess the prevalence and possible association between COVID-19 and herpes simplex virus (HSV) infection.

**Patients and methods:** A 9-item questionnaire was sent to 120 PCR-confirmed COVID-19 patients with a response rate of 66.67%. This cross-sectional observational study included 80 patients with mild to moderate COVID-19 infection who did not require hospitalization or steroid therapy.

**Results:** One or more HSV infections were observed in 28 patients (35%) with COVID-19 infection, including 10 males (35.7%) and 18 females (64.29%). Of the 28 patients, fever was reported in 17 patients (75%) during COVID-19. Most of the respondents (78%) described a single HSV reactivation, 14.29% had 2 attacks, and 7.14% experienced 3 attacks. Compared to previous non-COVID-19 related HSV reactivation, the COVID-19 related attacks were more severe in 12 patients (42.85%), equally severe in 5 patients (17.85%) and less severe in 1 patient (3.57%). Interestingly, 10 patients (35.71%) developed an initial symptomatic HSV attack during COVID-19 infection.

**Conclusions:** This study demonstrated a possible association between COVID-19 infection and primary HSV infection and/or reactivation. The COVID-19 direct neuronal effect in addition to COVID-19 related psychological stress, fever and immunological dysregulation could play a potential role.

## Introduction

For about a year now, the world has been under the grip of the COVID-19 pandemic. More than 183 million cases and approximately 4 million deaths have been reported globally (1).

COVID-19 is caused by severe acute respiratory syndrome coronavirus 2 (SARS CoV2), a member of the Coronaviridae family. In the 21st century, two other major respiratory disease outbreaks were caused by closely related coronaviruses, including severe acute respiratory syndrome (SARS) and Middle East respiratory syndrome (MERS) (2).

Although respiratory manifestations seem to be predominant, an increasing number of COVID-19 related cutaneous manifestations have been reported, including maculopapular, urticarial, purpuric, chilblain-like and vesicular eruptions. Other less commonly reported lesions include livedo reticularis-like, erythema multiforme-like, papulosquamous, necrotic lesions and gangrene (3) (4) (5).

Herpes simplex virus (HSV), an enveloped DNA virus belonging to the *Herpesviridae* family, is a ubiquitous pathogen that commonly infects humans. It has been estimated that two-thirds of the population under 50 years of age are infected with herpes simplex virus type 1 (6). After primary infection, HSV can establish a life-long latent infection in the sensory ganglia via retrograde transport through peripheral neurons. The virus can be reactivated periodically in response to various stimuli including psychological stress, fever, sunlight, hormonal imbalance, immunosuppression and surgical resection (7) (8). When HSV-1 infection is symptomatic, the most common clinical manifestations are gingivostomatitis and pharyngitis in children and adults, respectively, whereas Herpes labialis “cold sore” represents the most frequent sign of HSV-1 reactivation (9) (10). Other less frequent cutaneous manifestations of HSV-1 include herpetic whitlow, erythema multiform, eczema herpeticum, and herpes gladiatorum (11) (12) (13) (14).

## Patients and methods

A cross-sectional observational study was conducted at Baghdad Teaching Hospital, where data were obtained from an online survey conducted between August and October 2020. Eligibility criteria were as follows: any age, PCR-confirmed mild to moderate COVID-19 infection, not admitted to the RCU or required steroid therapy. A 9-item online questionnaire was created and sent by email to 120 patients with PCR-confirmed mild to moderate COVID-19. The survey included a range of measures to examine the prevalence, severity and timing of herpes simplex during COVID-19 infection. Participants reported their age, sex, COVID-19 status, history and severity of past HSV infection, history and severity of HSV during COVID-19, history of fever and HSV infection timing during COVID-19. The diagnosis of herpes labialis was based on the clinical features.

## Results

The questionnaire was completed by 80 out of 120 patients who received an online survey with a response rate of 66.67%. The mean age and SD were 33.87±9.46 years. Twenty-eight patients (35%) reported single or multiple episodes of HSV reactivation (Table 1). The majority of patients (n=24, 78.57%) reported a single attack of HSV reactivation during their COVID-19 infection, while 14.29% and 7.14% experienced two or more attacks, respectively (Figure 1). Of the 28 patients, 18 (46.29%) reported recurrent attacks of HSV reactivation before COVID-19 infection. Compared to previous attacks, COVID-19 related HSV reactivation was more severe, equally severe or less severe in 42.86%, 17.68% and 3.57%, respectively. The onset of HSV infection varies among individuals after the onset of COVID-19. The mean time of onset of the first HSV infection was 8+6.48 days while the onset on the second and third attacks were 11+3.44 and 22+3.53 days, respectively (Table 2).

**Table 1:** Frequency distribution of the COVID-19 related HSV infection, fever and previous HSV infection among included patients

|  | Yes | No |
| --- | --- | --- |
| **Did the patient get an attack of herpes simplex during the COVID-19 infection?** | 28 (35%)<br>• Male 10 (35.71%)<br>• Female 18 (64.29%) | 52 (65%) |
| **Did the patient get a fever during the attack?** | 21 (75%) | 7 (25%) |
| **Does the patient have recurrent attacks of HSV before COVID-19?** | 18 (64.29%) | 10 (35.71%) |
| **Was the COVID-19 related HSV the first symptomatic HSV attack?** | 10 (35.71%) | 18 (64.29%) |

**Figure 1:**
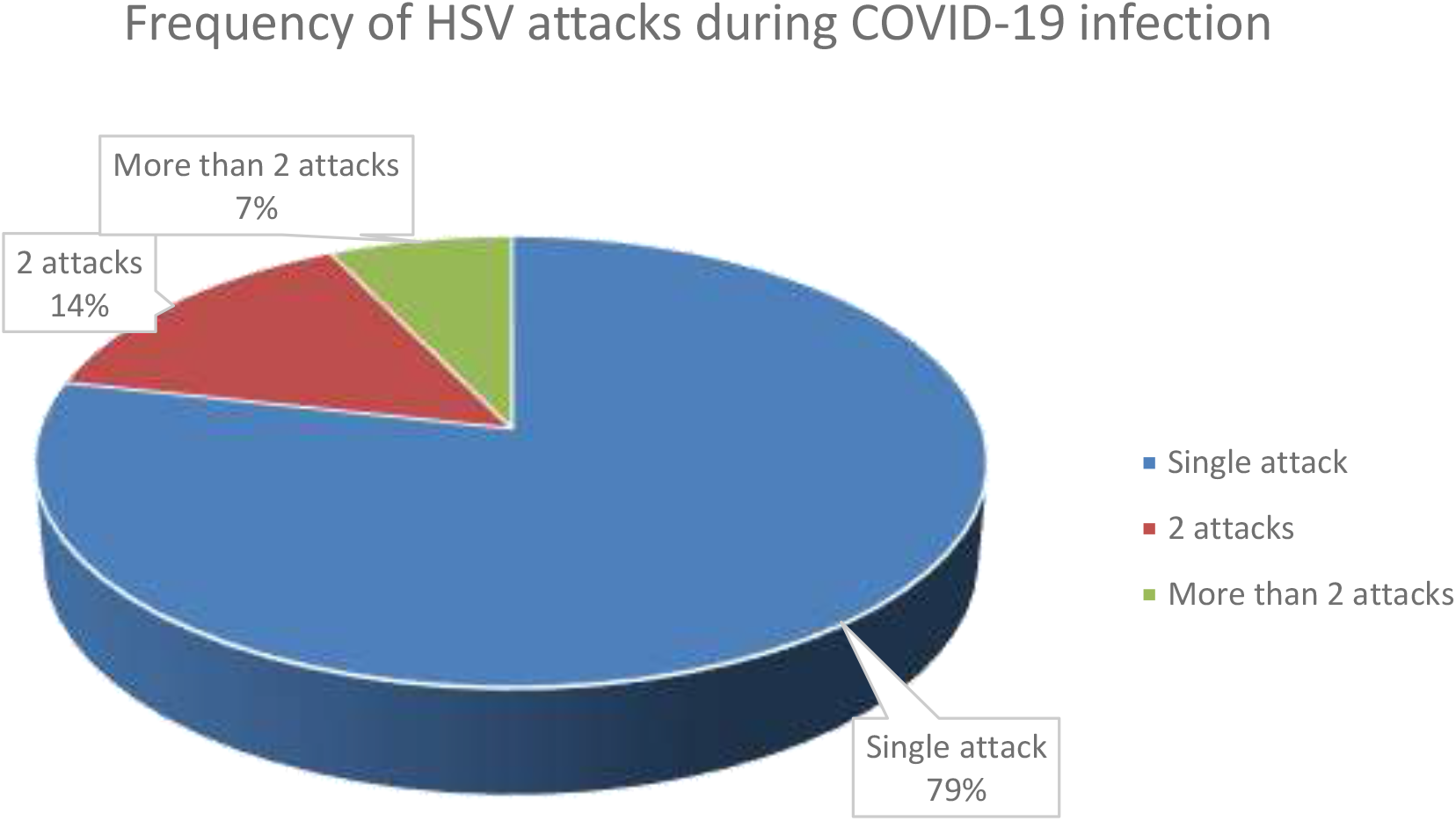
Frequency of HSV infection among COVID-10 patients.

**Table 2:** The mean time of HSV onset after acquiring COVID-19

| **Timing of HSV reactivation (days)** | **No.** | **Mean** | **SD** |
| --- | --- | --- | --- |
| **First attack** | 28 | 8 | 6.48 |
| **Second attack** | 6 | 11 | 3.44 |
| **Third attack** | 2 | 22 | 3.53 |

## Discussion

Depending on the host cell type, HSV can produce lytic and latent lytic stages, where the lytic stage can occur in various tissue types, while the latent stage tends to occur in neuronal tissues (15). The lytic stage can result from viral replication in the epithelial cells of the oral and genital mucosa, eye or skin producing labial herpes (cold sores), genital herpes, herpetic keratitis and herpetic whitlow or eczema herpeticum, respectively. HSV is a neurotropic virus that can establish latency in the neuronal dendrites of the sensory ganglia that supply primary epithelial tissues (16). The immune system plays a vital role in controlling HSV replication, driving the virus into a latency state for prolonged periods (17).

A total of 28 patients with PCR-confirmed COVID-19 (35%) reported single or multiple attacks of HSV infection (Figure2), including 18 females (64.29%) and 10 males (35.71%) with a mean age of 38.4 years and a standard deviation of 13.64. The number of females surpassed that of males; as the rate of HSV seropositivity is slightly higher in females aged between 15 and 39 years than in males (18) (19).

**Figure 2:**
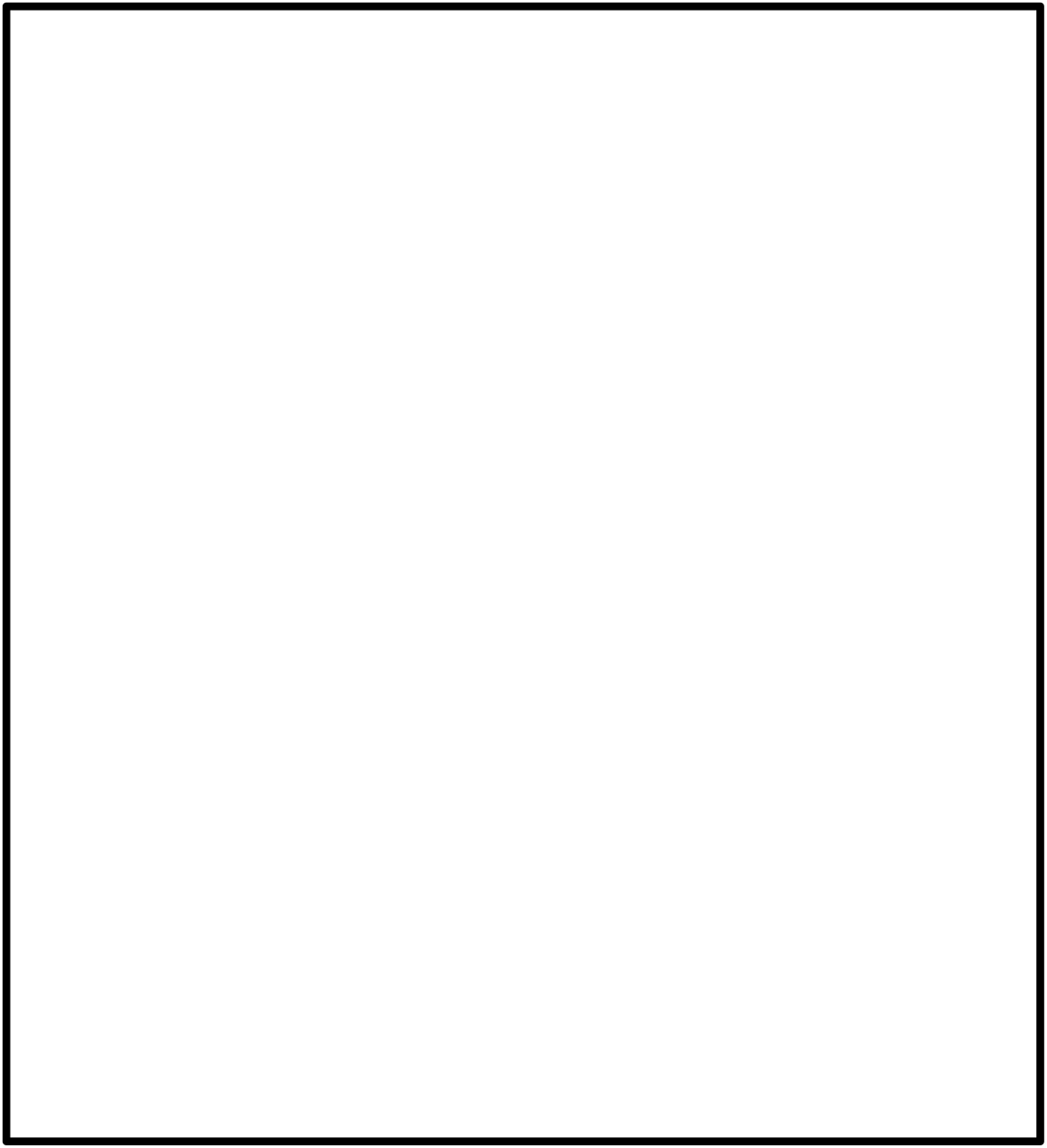
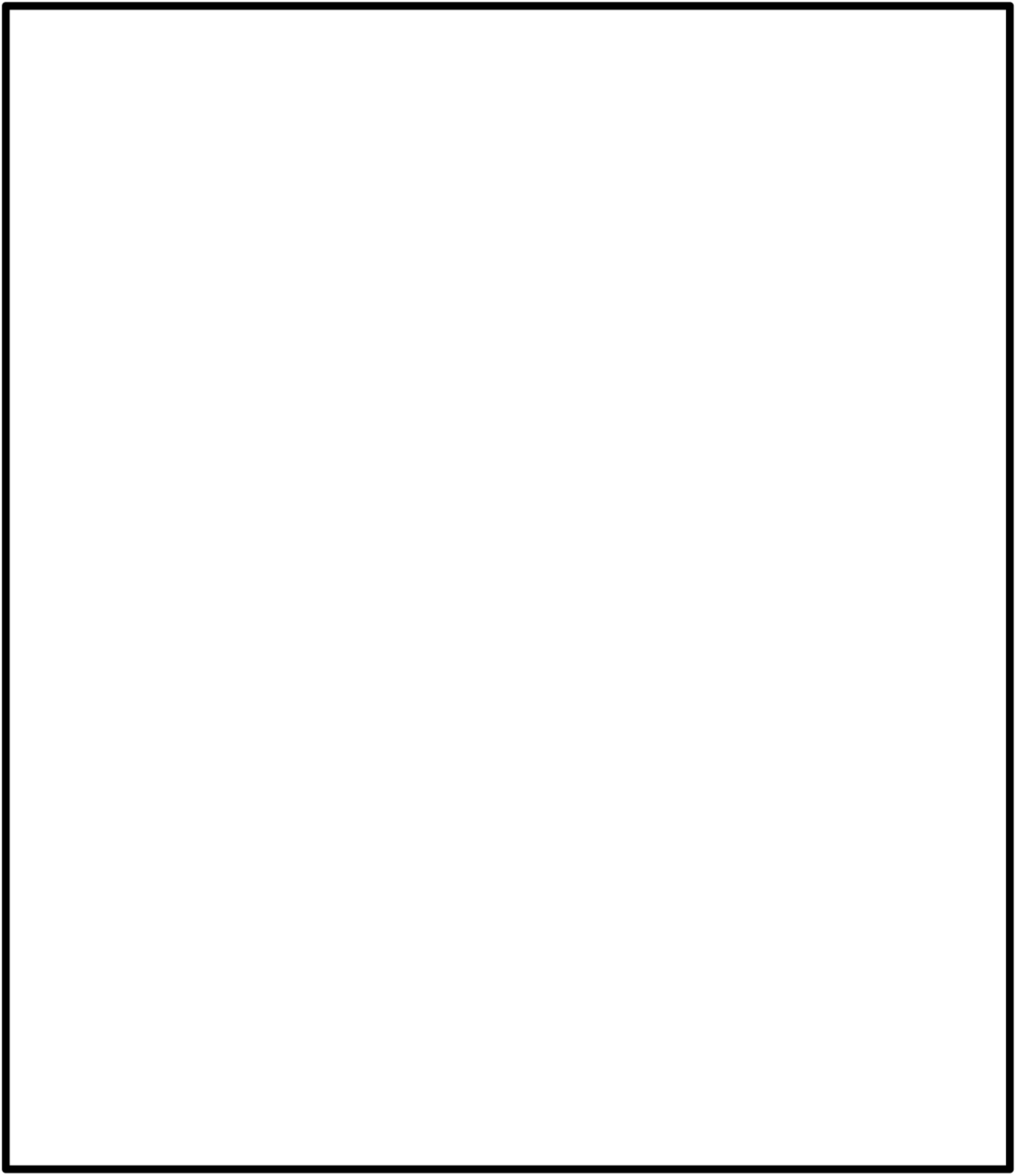
Two examples of HSV infection (herpes labials) in COVID-19 patients. ***Facial Photos were removed according to Medrxiv policy***

Four patho-mechanisms were proposed to explain the higher frequency of herpes reactivation during COVID19 (Figure 3):

**Figure 3.**
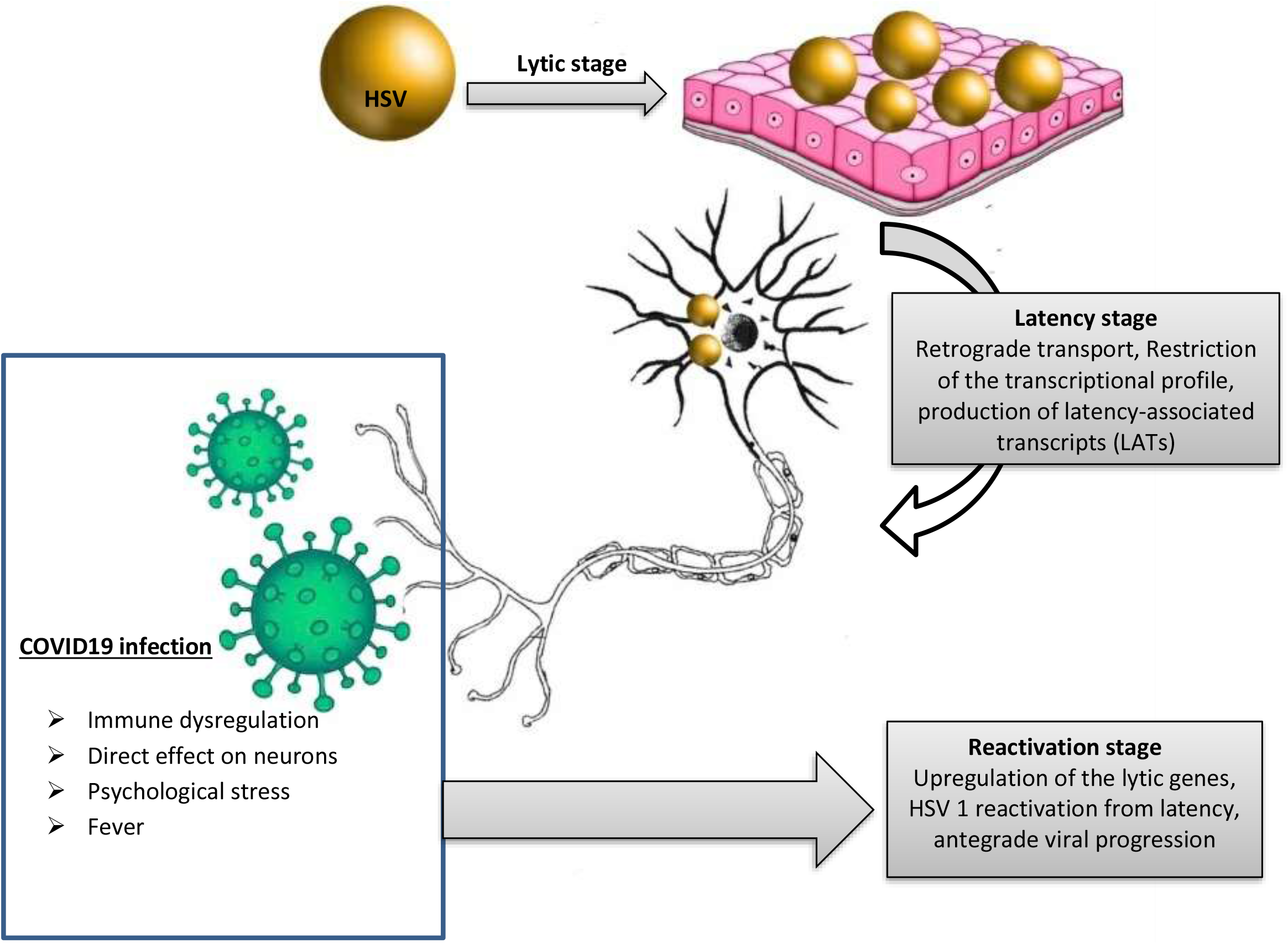
Proposed patho-mechanisms of HSV reactivation during COVID-19 infection. After primary infection, HSV can remain in the latent stage for several years. COVID-19 related immune dysregulation, fever, stress and the direct neuronal effect of the virus can result in upregulation of HSV lytic genes, reactivation of the latent virus and viral replication.

### 1- COVID-19 related immune dysregulation

COVID19 is associated with unique immune dysregulation, including CD4 cell and NK cell cytopenias and sustained inflammatory interleukins stimulation such as interleukin-6 (IL-6) and tumor necrosis factor-α (TNF-α) (20). Immune dysregulation causes HSV-1 to burst from latency and travel anterogradely to epithelial surfaces, where viral replication and lytic stages occur (21). Interestingly, a potential relationship between the cytokine interleukin (IL)-6, which is overrepresented in COVID19 patients (22) (23) and reactivation of herpes simplex virus has been proposed (24). Compared to mice receiving control antibodies, HSV-latently infected mice injected with neutralizing anti-IL-6 antibodies show a lower frequency of virus reactivation (25) (26).

### 2- Direct effect of COVID19 on the neurons

COVID19 has a potential neurotropic mechanism that explains the neurological manifestations associated with COVID-19 like loss of taste and smell, headache, dizziness, meningitis, cerebrovascular disease and acute Guillain–Barré syndrome. SARS-CoV-2 has an affinity to bind to the angiotensin-converting enzyme 2 (ACE2) receptor; hence, cells that express ACE2 like neurons and glial cells are vulnerable to SARS-CoV-2 infection (27) (28).

### 3- COVID-19 related psychological stress

COVID19 is associated with significant psychological and physical stress. It has been postulated that stress impairs the cytotoxic T cell surveillance of latently infected neurons, resulting in replication and activation of latent viruses (29) (30) (31). Other hypotheses include stress-related relapse of catecholamines and glucocorticoids (stress hormones) and their direct and indirect effects on HSV reactivation (32).

### 4- COVID-19 related fever

Seventy-five percent of patients with HSV in the survey reported fever. Fever is a strongly linked environmental trigger of human HSV reactivation either through a direct effect on latently infected neurons and/or the secretion of pyrogenic cytokines, including IL-6 (33) (34).

Le Balc’h et al. suggested that COVID-19 infection could be a risk factor for *Herpesviridae* reactivation and subsequent pulmonary infection in patients with COVID-19 severe acute respiratory distress syndrome (35).

Elsaie et al described two cases of varicella-zoster virus (VZV, another member of the Herpesviridae family) during COVID-19 infection. After primary infection, VZV remains dormant in neuronal tissues and is reactivated in a similar fashion to HSV (36).

## Conclusion

COVID-19 related immune dysregulation, psychological stress, fever and direct neuronal effects might play a role in the activation of different cellular processes that result in increased HSV lytic gene expression and reactivation of the virus. Herpes simplex reactivation in patients with new respiratory symptoms could serve as a new clue of the emerging COVID-19 infection.

## Data Availability

N/A

## Limitation of the study

Recall bias is expected, and further prospective or retrospective case-control studies are advisable.

